# Triad entanglement of estrogen-sulfotransferase (SULT1E1), NFκβ and Nrf-2 confers matrix-metalloprotease (MMP 2/9) action in breast carcinogenesis

**DOI:** 10.1101/2020.04.17.20068957

**Authors:** Aarifa Nazmeen, Sayantani Maiti, Smarajit Maiti

## Abstract

Estrogen (E2) is one of the most important signaling molecules that control cell-differentiation/early-embryogenesis/organogenesis in gender-independent manner. Nevertheless, during adolescence/adulthood it influences female reproductive-functions by delicate cellular proliferative-events via nongenomic (cellular-signaling)/genomic (transcriptional-signaling) pathways to recruit a number of genes/proteins. In case of post-menopausal-women high E2 may initiates tumors in breast/gynaecological-tissues. Impired estrogenic signaling may be the results from abnormal redox-regulations of estrogen-metabolizing-enzyme estrogen-sulfotransferase(SULT1E1), transcriptional-factors NFκβ, Nrf-2 and Matrixmetalloproteases (specially MMP 2/9) in the breast-tumor. Here, tumor and its surrounding tissues were obtained from the district-hospital. Intracellular redox-environment of tumors was screened with some in vitro-studies. RT-PCR for SULT1E1 expression and MMP 2/9-zymogram were conducted in lasoprazole (Nrf-2 inducer) or dexamethasone (SULT1E1 inducer) treted rat liver tissues. Immunohistochemistry was performed to analyze SULT1E1/NFκβ localization and MMP 2/9-zymogram in human breast-cancer versus its surrounding tissues. It can be hypothesized that transcription-factors (NFκβ/Nrf-2) imposes effect on MMPs expressions resulting in significant impacts on metastatic transition of breast-cancer. Breast tumor reveals higher (vs surrounding-tissue) expression/immunolocalization of NFκB/SULT1E1 paralleling to our previous finding of Nrf-2 induction. The relation between Nrf2/NFκB is determined by oxidative-stress and by CBP recruitment of HDAC3. Further, this relation is a determinant of MMP-regulations and SULT1E1-mediated E2 levels. Adaptively, augmented Nrf-2 may induce SULT1E1 resulting in lower active-estrogen. The triad regulations of NFκβ, SULT1E1 and Nrf2 are proposed here to execute MMPs function in the severity of human breast-carcinogenesis. Therapeutically this triad system may be effectively targeted for breast cancer treatment. Further studies are necessary in this regard.

## INTRODUCTION

Estrogen is an important regulator of growth and differentiation in various tissues, such as the liver, reproductive tracts, central nervous system (CNS), and mammary gland (Nilsson et al., 2004). Estrogen by tradition has been associated with female reproduction. However, the importance of estrogen and other hormones in metabolism and functions like growth has been established later (Ropero et al, 2008). In post menopausal women high estradiol level may initiates tumors in breast/gynaecological-tissues which may be correlated to several tumerogenic marker cum biochemical regulator molecules (Nazmeen et al. 2017). Developing breast cancer involves altered genetic factors, and their mutation as well as long term exposure to estrogen during the entire lifetime. The molecular mechanisms responsible for estrogen influenced/induced breast tumor formation are still not clear. While, estrogen receptor α (ERα) is decisively involved, non-receptor mediated effects of estradiol (*E*_2_) may also play an important role in breast tumor development (Nazmeen and Maiti, 2018). Studies reveal that in ERα knockout (ERKO) mice bearing the Wnt-1 oncogene, there is an ERα independent role of *E*_2_ on mammary tumor development. Exogenous *E*_2_ accelerated tumor formation in a dose-dependent fashion at early follicular and mid luteal phase in ERKO/Wnt-1 animals. Lowering endogenous E2 levels by oophorectomy or an aromatase inhibitor (AI) in intact ERKO/Wnt-1 animals delayed the process of tumorigenesis. This can be a strong evidence for an ER-independent effect of estrogen on tumor development (Wei et al., 2010).

Metastasis is a marker of disease severity and associated with negative prognostic effects. MMPs are largely involved in the metastatic event. MMPs activates a number of tumorigenic process in breast cancer. These events can directly facilitate progression of cancer by degrading basement membrane that allows cancer cells to invade the surrounding tissue and enter blood stream also. MMPs may also impose direct effect on the tumor cells leading to release molecular factors that promote growth and or suppress apoptosis (Gialeli et al., 2011).

Cancer cells exhibit an accelerated metabolism and demand or produce higher amount of ROS to manage their elevated proliferation rate. Cancer cells adapt diverse ways of developing ROS resistance, including the implementation of alternative pathways. The adaptation of alternative pathways can avoid accumulation of large amount of ROS without compromising the energy supply required by cancer cells (Maiti and Nazmeen, 2019). The adaptive processes include leading the glycolytic pathway into the pentose phosphate pathway (PPP) along with the generation of lactate and stop the aerobic respiration in the mitochondria (Sosa et al., 2013). There is strong relation between ROS level and ROS sensitive different gene expression that is related to the cell regulatory processes. ROS mediated inflammatory processes that keenly involved in the breast cancer pathogenesis. In this regard inflammatory transcription factor NFκβ is of great importance. It agonizes the inflammatory immunological responses in ROS dependent manner (Nazmeen et. al., 2020). Metabolism in different antigen presenting cells createshyper metabolic state resulting in chronic hypoxic conditions. Nevertheless, the increase of ROS initiates higher expressionof NFκβand HIF-α. Nrf2 mediated pathways are also utilized in cancer cells in order to control the ROS levels. Nrf2 is known to be responsible for induction of the antioxidant enzymes, which may be an inhibitor of oxidative stress induced tumorigenesis. Oxidative stress or ROS accounts for several signalling pathways, which helps cancer cell thrive better. Hence, reduced Nrf2 in such cases may be a threat to the patients. Whereas, an elevated Nrf2 in certain cases of breast cancer may allow better survival of the cancer cell, via creating a balance between oxidants and antioxidants. Thus, upregulation or downregulation of Nrf2 favorably depends on the disease stage and environment. In this paper, we have reviewed the concerted involvement of SULT1E1, Nrf2 and NFκβin breast cancer and their crosstalk.

## Material and Methodology

### Ethical clearance and fulfillments of other regulatory affairs

An informed consent was obtained from all the pateints included in this study. This study was conducted according to the National Institutes of Health, USA guidelines and all the instituitional ethical guidelines and regulations were strictly followed during the investigation. The experiments in this study were given approval by institutional ethics committee (oist/EC/hu/bt/16/).

Female Wistar rats were purchased from a farm house which is a Government accredited [CPCSEA-Committee for the Purpose of Control and Supervision of Experiments on Animals: Reg. no. 1A2A/PO/BT/S/15/CPCSEA ⍰(http://cpcsea.nic.in/Auth/index.aspx)] organization.

### Inclusion and exclusion criteria of the breast cancer pateints

Patients only with breast carcinoma undergoing mastectomy were included. Patients suffering from any other disease except breast carcinoma were all excluded. Women with Pregnancy, menstruation, HIV, HCV, HPV, and Hepatitis B were all excluded.

### Sample collection

The breast tumor samples were collected from local District Medical College and Hospital. Tumors were clinically diagnosed according to the TNM [The extent of the tumor (T), the extent of spread to the lymph nodes (N), and the presence of metastasis (M)] classification system. Surrounding tissues and corresponding tumor samples were collected in separate aliquots exactly after surgery and stored at −⍰20 °C. A partof both tumor and surrounding tissue were stored in formalin immunohistochemistry. The antibodies against NFκβ(PB9149) were purchased from BOSTER BIOLOGICAL TECHNOLOGY, CO LTD, 3942B Valley Avenue, Pleasanton, CA 94566.

### Cytosol preparation

A 30% w/v homogenate was prepared with the breast tumor and the corresponding surrounding tissues in ice-cold phosphate buffer (0.1 mol/l, pH 7.4). The supernatant (cytosol) was collected after centrifugation at 10,000 rpm at 4 °C for 30 min and stored at −lil20 °C in separate aliquots for assays.

### Immunohistochemistry of NFκB and SULT1E1 in human breast-tumor vs surrounding tissues

Paraffin blocks are prepared according to the standard method; an automated cryostat slicing machine (Leica Bio systems) was utilized to serially section the blocks at 5 μM. The deparaffinisation was done by baking at 60 °C followed by treatment with xylene, alcohol downgradation and treatment with water. The immunohistochemistry technique was done according to the standard method. The method included washing with PBST + 1 % casein for 10 min, sections were then incubated with 5 % casein for 30 min which prevented non-specific binding and incubated in primary antibody against NFκβovernight, washed with 1% PBST and then incubated in secondary antibody for 1 h. Slides were washed with 1% PBST then by water and stained with DAB for 3 min and then washed with water. Slides were mounted with mounting medium and observed under a microscope (Nikon, Eclipse LV100, magnification 20×) to study the NFκβ expression and localization.

#### In-vitro induction of SULT1E1 rat hepatocytes

### Preparation of single cell suspension of rat hepatocyte

50 g of rat liver tissue was scrapped through nylon mesh in buffer (DBSS). Centrifugation of the homogenate was done at 120×*g*. The pellet was washed with L-15 media containing 1500 mg/l D-glucose, 1% penicillin and streptomycin, 20% FBS, 1% Glutamine. After washing the Cells were kept in 10 ml of L-15 media. 650 µl of media containing cell was added to each petri plate comprising of 9350 µl of L-15 media (1500 mg/l D-glucose, 1% penicillin and streptomycin, 20% FBS, 1% Glutamine).

### Lansoprazole/dexamethasone treatment to rat hepatocytes

Two drugs, Dexamethasone and Lansoprazole were used to treat the cell suspension. Dexamethasone was added to petri plate at a concentration of 100 µM. 25 mg of Lansoprazole was added to petri plate containing cell suspension. The culture was incubated for 72 h in an incubator at 37 °C.

### MMP 2/9 zymographic analysis in breast tumor and experimental rat-hepatocyte model

The MMP gel assay was performed by following a standard method. A 1 % type B gelatin solution is prepared in water, 8 % resolving gel is prepared containing 1% gelatin solution, samples were prepared in MMP dye and loaded with the same dye. Gel was allowed to run at 110V for 60 minute. After removing the gel was activated by washing 3 times with gel washing buffer containing Triton X-100 and other reagents. The gel was then incubated in zymographic gel development buffer pH 7.4 at 37°C for 42 hours. Gel were then stained in amide black or coommasie blue containing solution for 1 hour at room temperature in a shaker. After an hour the gels were then destained with solution I for 15 to 30 minutes followed by destaining solution II for 3 to 5 hours until clear bands appeared. Destaining solution was then washed out with sterile water for 30 minutes and then incubated in a gel preservating solution for 15 minutes. Immediately after 15 minutes the gels were scanned in a digital scanner.

### RT-PCR of SULT1E1 in lansoprazole/dexamethasone treated rat hepatocytes

RNA was isolated from cells treated with dexamethasone and lanzoprazole. 1 μg of whole RNA was utilized to perform reverse transcription PCR was using Qiagen one step RT-PCR kit following the instructions provided in the kit.

## Result

### Higher immunolocalization of NFκB and SULT1E1 in human breast-tumor

NFκB and SULT1E1 are highly expressed in the tumor tissues of breast cancer patients. The stromal regions were darkly stained both for SULT1E1 and NFκB in tumors whereas the surrounding tissue also showed expression but lesser than the tumor. The surrounding tissue has less cellular region than tumors, and possess more of adipose tissue visible as white or light brown patches (Fig-1).

**Fig-1:**
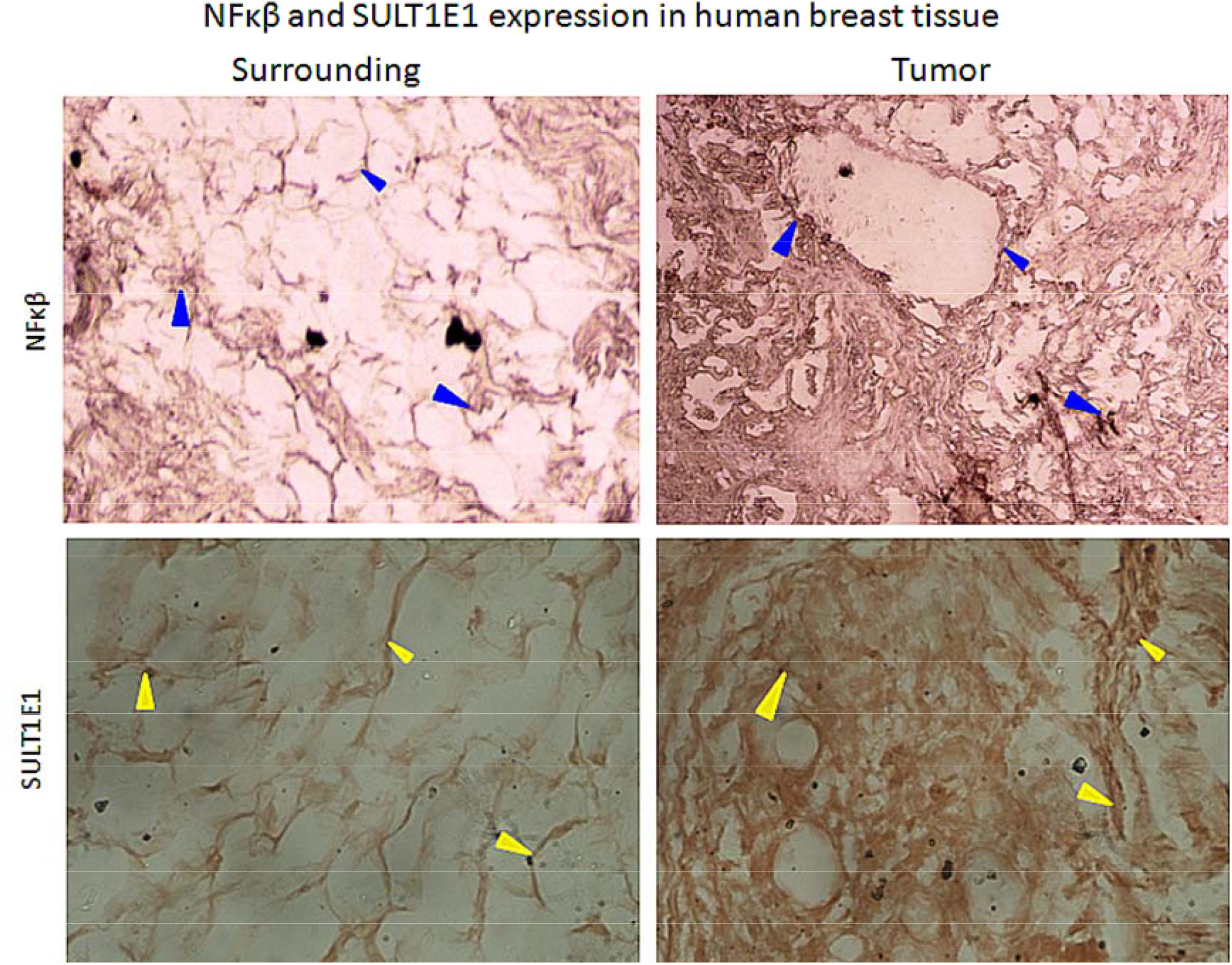
Immunohistochemistry of breast tumor and its corresponding surrounding tissue. Tumor shows an increased expression of SULT1E1 and NFκB as compared to their corresponding surrounding tissue.

### Higher SULT1E1 mRNA expression in lansoprazole/dexamethasone treated rat hepatocytes

Dexamethasone seems to be a strong inducer of SULT1E1 as compared to the control and lansoprazole. Dexamethasone is a known inhibitor of inflammatory cytokines. Lansoprazole a known inducer of Nrf2 also showed induction of SULT1E1 greater than control but less than dexamethasone. Both inflammatory cytokines and Nrf2 has evident roles in the induction of SULT1E1 and MMP (Fig-2c).

**Figure-2:**
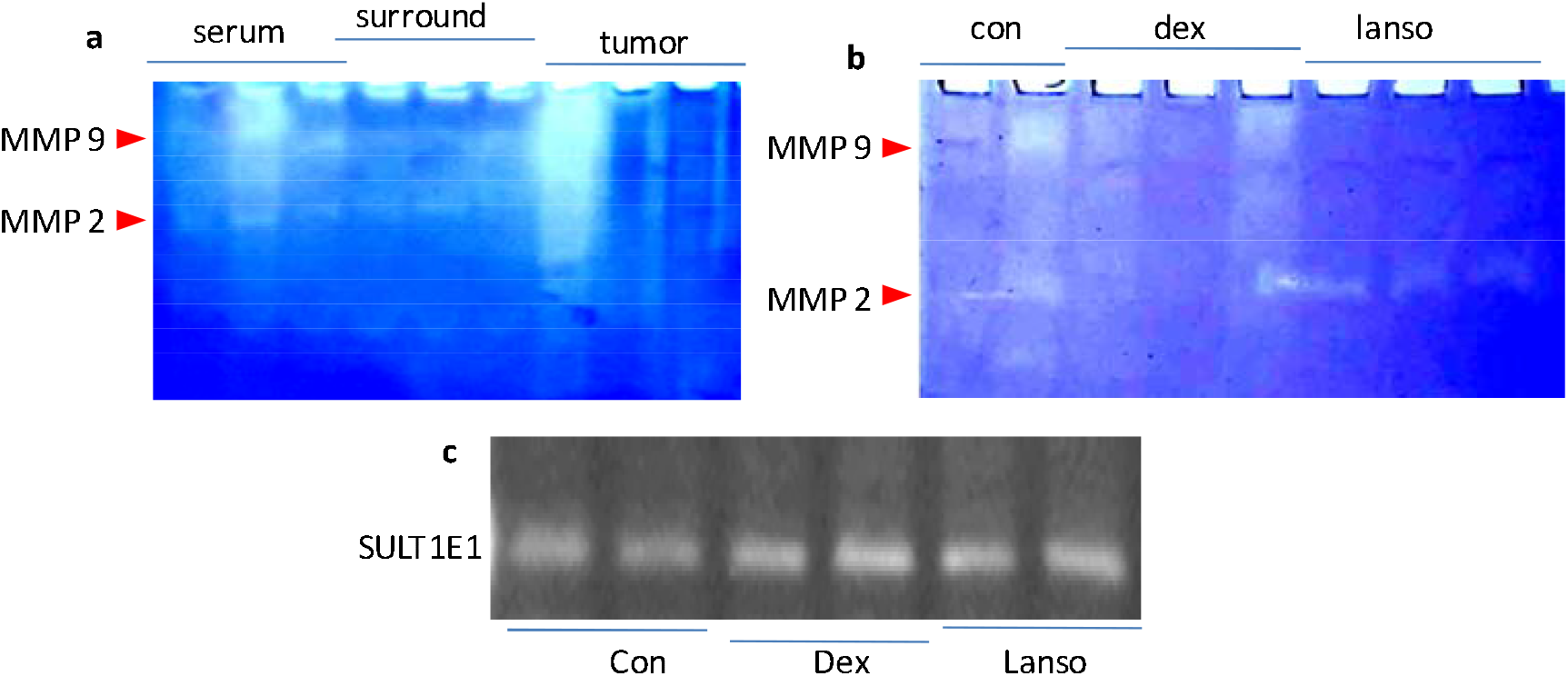
**a-** MMP activity in serum, surrounding tissue and the tumour tissue of breast cancer pateints. **b-** MMP activity in the invitro cell culture treated with dexamethasone and lansoprazole. **C-** Expression of SULT1E1 mRNA in dexamethasone and lansoprazole treated in vitro cell culture

### Variable MMP 2/9 zymographic signal in breast tumor and experimental rat-hepatocyte model

The serum and the surrounding tissue shows more MMP-9 activity than MMP-2. The tumour tissue shows no or least MMP-9 and MMP-2 except the first tumor sample (Fig-2a), MMP2 was expressed and highly active in the lanzoprazole group as compared to the dexamethasone, whereas, there is no MMP activity and some activity was noticed in the control group. This result clearly shows mmore inhibitory impact of dexamethasone on MMPs than that of Lansoprazole (Fig-2b). Figure 3 explains the summary of NFκB, Nrf-2 and SULT1E1 relationship. Proinfllamatory regulations of NFκB can be synergized with oxydatively regulated Nrf-2 action.

**Figure-3:**
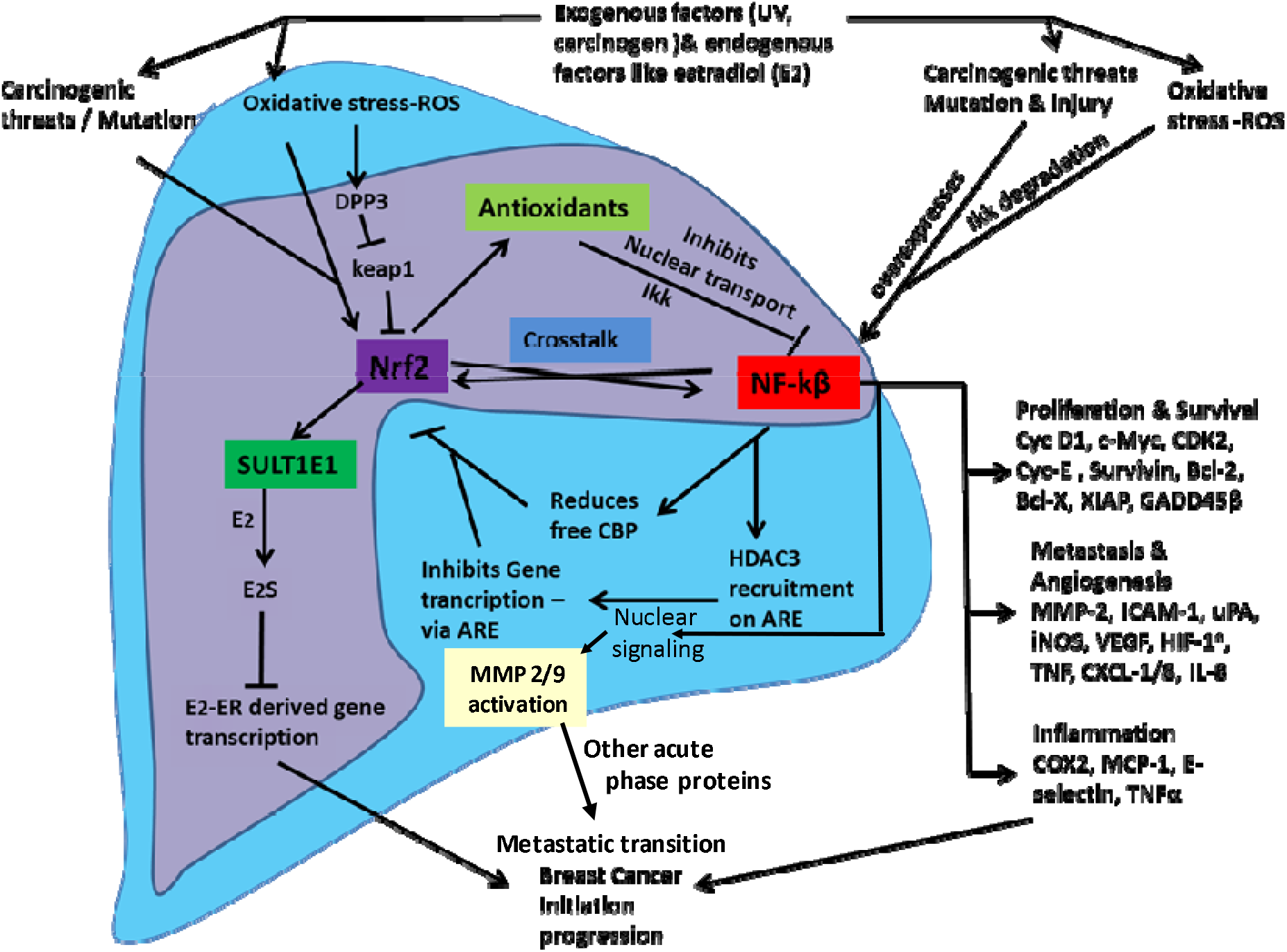
Triad role of SULT1E1, NFkB and Nrf-2 in breast cancer.The entire mechanism possibly showcases the scenario in case of established breast carcinoma. Based on the current studies, we have made an attempt to sort out or cleave this mechanism in two different pathways which can be utilized for prevention and treatment. 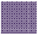 The purple region showcases the pathway which can be targeted to prevent disease initiation. 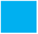 The blue region showcases the pathway to be targeted in order to treat the disease or reduce disease severity. This pathway can also be utilised to make cancer cells more susceptible to ROS inducing drugs.

Both are independently and additively related to the tumerigenesis. But more response of Nrf-2 related SULT1E1 expression midht be a better determinant of E2 level in serum and tumor tissues of breast cancer patients. The triangular function of these moleculae finally impact on MMP activities and severities in breast carcinogenesis

## Discussion

Matrix metalloproteinases (MMPs) are zinc-dependent proteases. MMP-2 and MMP-9 are vital metaloproteinases (Chia et al., 2014: Stoeltzing et al., 2003). MMP-2 and MMP-9 are mainly secreted in the form of zymogens by tumor cells and stromal cells. MMPs are activated via hydrolysis and are then ready to degrade basement membrane (BM) type IV collagen, which makes basement membrane less able to impede tumor cell movement (Zhang et al., 2014). In the current study, consistent MMP-activities in the surrounding tissues compared to the patients tumor tusses and serum suggests that there are some transition in molecular signaling. This has been further shown in terms of NFκBand SULT1E1 expressions in the tumor tissues. This suggests that disease severity is multi factor dependant.

Evidences support a role for MMPs in the initiation of carcinogenic process (Duffy et al., 2000) circulating MMPs may increase among women prior to the development of clinically detectable breast cancer. Research has established that MMP-2 and MMP-9 have key roles in degrading extracellular matrices to promoting tumor metastasis and invasion (Iochmann et al., 2009: Safrane et al., 2009).

MMP2 is thought to facilitate tumor development via growth factors processing (Klein et al., 2010: Levi et al., 1996) influencing inflammatory markers and stimulation of angiogenesis (Fang et al., 2000: Egeblad et al., 2002). A few earlier studies also indicates that MMP2 expression and activity may partially be regulated by estrogen (Nilsson et al., 2007: Philips et al., 2004), a hormone which proves to have well-established roles in promoting breast cancer initiation and progression (Bernstein et al., 1993). An elevated estrogen has also been noticed in tumors compared to surrounding in earlier studies from our lab (Nazmeen et al., 2020). The current study shows an elevated SULT1E1 (Fig-1) that may lead to reduced E2, If SULT1E1 is in active state. On the other hand an elevated oxidative stress leads to inactivation of SULT1E1 enzyme (Maiti et al., 2007), despite of an increased expression. Preinvasive breast cancer shows lower stromal aromatase expression and high-grade DCIS shows lower SULT1E1 levels. This suggests that intratumoral estradiol (E2) modulating enzymes results in lower local E2 levels in the early stages of breast tumorigenesis (Hudelist et al., 2007) but gradually E2 elevates with the upgradation of the disease and MMP is also associated with the high grade metastatic cancers, thus may be sharing some connection. In the cases of lymph node metastases estrogen receptor α and stromal aromatase expression were correlated

Potential connection between MMP2 and estrogen are incompletely understood, and needs an in depth research since in vitro and human studies have shown conflicting results(Abdallah et al., 2007: Katayama et al., 2009: Pitteri et al., 2009). The relationship between circulating MMP2 and risk of breast cancer is inconsistent as well as limited. Several studies also exist that do not support the utility of plasma MMP2 as a biomarker to detect breast cancer risk, although MMP2 warrants further research as a potential early indicator of tumor aggressiveness. In the current study the serum sample and the surrounding tissue shows more MMP9 than MMP2 (Fig-2a). The tumour tissue shows no or least MMP9 and MMP2 except tumor sample no 16. MMP2 was expressed and active in the lanzoprazole group as compared to the dexamethasone (DEX), where there is no MMP activity and slight activity was seen in the control group (Fig-2b). DEX is a strong inducer of SULT1E1 (via inhibition of inflammatory cytokines) whereas lanzoprazole (via Nrf2 induction) also showed induction of SULT1E1 greater than control but less than DEX (Fig-2b). Studies suggests that anti-proliferative effect of some progestin such as Nomegestrol acetate andMedrogestone in breast tissue relay via stimulation of SULT1E1 and inhibition of STS pathway (Chetrite et al., 1999). So, both DEX and lanzoprazole may be effective antiproliferators in breast cancer via SULT1E1 induction.

The proinflammatory transcription factor nuclear factor-kappa B (NFκB), is mostly found activated in breast cancer. NFκB accelerates the disease severity leading to high grade or late-stage tumor phenotype via facilitating hormone-independence of the breast cancer (Wang et al., 2015). Inhibition of NFκB by Bay-11-7082 resulted in the reduction of CD44 expression. CD44, a cell surface glycoprotein, is involved in many cellular processes, including cell adhesion, migration and proliferation (Smith et al., 2014).The pathways activating or activated by NFκB are highly involved in the process of breast cancer progression targeting some specific progenitor cells (Shostak et al., 2011).Decontrolled NFκB activation causes persistent nuclear localization of p50, p52, p65, cRel and RelB proteins. Nuclear localization of these proteins breaks the coordination between cell proliferation and death through the up regulation of anti-apoptotic proteins (Karin M et al., 2002). NFκB modulators are being used to suppress cancer metastasis (Liu B et al., 2015).

Cell culture treated with lanzoprazole induced MMP2 activity which abides the earlier study that lanzoprazole administration increases MMP-2 expression, in a Rat Model of Acetic Acid-Induced Gastric Ulcer (Kobayashi et al., 2010). In this study both NFKB and SULT1E1 are highly expressed with no MMP in tumors (Fig-2a), and this results seems to be comparable with DEX results which induces SULT1E1 (2c) but there is no MMP in this pathway of SULT1E1 induction. There seems to have something in common between these two results. Lanzoprazole also induces expression of SULT1E1 (Fig-2c) via Nrf2 induction and along with that it presents MMP expression also.Nrf2 is a transcription factor of the leucine zipper family. Nrf2 regulates antioxidant enzymes, conjugation of electrophile, homeostasis of glutathione, production of reducing equivalents, and proteasome function etc (Hayes & McMahon, 2009).Cancer cell proliferation is inhibited by lutein via alleviating oxidative injury. Nrf2 /antioxidant response element (ARE) and blocking of NFκB pathways together mediates anti proliferative activity, but the mechanism is not well understood (Chang et al., 2017).Reports reveal that cancer stem cells (CSCs) maintain reduced ROS levels by elevating the expression antioxidative enzymes and ROS-scavenging molecules, which directly favors the survival and chemoresistance of the CSCs. Brusatol an Nrf2 inhibitors can become novel chemotherapeutic drug order to combat refractory tumor initiating cancer stem cells CSCs (Wu et al., 2014).

Nrf2 is a transcription factor which regulates cellular redox status via endogenous antioxidant systems and simultaneously imposes anti-inflammatory activity (Ferrari et al., 2019). Earlier studies from our lab have shown that Nrf2 was also highly expressed in the tumor samples which may have some inhibitory impact on NFκB, which results in less or no MMP in tumors. Well this contradicts with the Lanzoprazole results which induce both Nrf2 and MMP2 (Fig-2a). HDAC inhibitor, trichostatin A enhances Nrf2 transcriptional ability via its acetylation and the expression of downstream targets, This finally protects against cartilage degradation via the reduction in matrix metalloproteinase (MMP)s and proinflammatory cytokines TNF-α, IL-1β, and IL-6 in osteoarthritis (Cai et al., 2015).HDAC inhibitors are known to have protective effects against cartilage degradation through mechanisms including Nrf2 activation and the inhibition of NFκB and MAPK (Khan and Haqqi, 2018).

Thus, it infers that NFκB, TNF-α, IL-1β, and IL-6 and HDAC are associated with the MMP expression and activity. Dexamethasone supresses IL-8 and the expression of NFκB in the nuclei and cytoplasm of mononuclear cells in tracheobronchial lavage fluid (TBLF) from premature neonates with respiratory distress (Zubair et al., 2006). Therefore dexamethasone is an established inhibitor of the NFκB mediated MMPs. LCIS patients showed MMPs expression in normal breast tissue and also in LCIS lesions in patients. However the expression in LCIS was not higher compared to the normal tissue. This suggests that MMP expression does not increase with the gain in a more proliferative phenotype (Nyante et al., 2019). Several reports on retrospective case-control studies have shown higher circulating MMP2 levels in patients than in age compared controls (Sheen et al., 2001: Patel et al. 2011), while some other studies have also observed no difference in MMP levels (Somiari et al., 2006: Katunina et al., 2011). Our results also showed an increased Circulating MMP2 and MMP9 along with increased MMP in the adjacent tissues (Fig-2a). Studies suggest that increased MMP-2 expression in carcinoma cells was found in stage I disease. Presence of Stromal MMP-2 was related to poor differentiation. These studies also support no or least expression of MMPs in Tumor tissue (Fig-2a). High MMP-9 expression in carcinoma cells was linked with small tumor size and lower recurrence (Johanna et al., 2004). The surrounding tisse showed increased MMP9 and MMP2 (Fig-2a) which is possible because as soon as secretd MMP is deliverd to its functional area and that is the extracellular matrix where it degrades Type-IV collagen constituents of the basement membrane separating tumors from surrounding tissue (Liotta et al., 1980). MMP is associated with extracellular matrix degradation, and MMP is associated metastasis, So Dexamethasone can be promising drug which can limit estrogen role by inducing SULT1E1 and along with that it prevent the disease to be proliferated and prevent metastatic transition of the disease by inhibiting NFκB other inflammatory cytokines and MMPs.

## Conclusions

The immunohistochemistry of breast tumor and its corresponding surrounding reveals that expression of NFκB is higher in tumor as compared to the surrounding (Fig-1). These tumors had higher value of E2 as per our earlier studies (report from hospital). Nrf2 may inhibit NFκB via antioxidants, whereas NFκB may inhibit Nrf2 either by reducing CBP or recruitment of HDAC3 on AREwhich inhibits gene transcription via ARE. On the other hand Nrf2 may induce SULT1E1 which reduces level of active estrogen. The entire mechanism possibly showcases the scenario in case of established breast carcinoma. A cancer cell perfectly finds out ways to balance proliferation and apoptosis via targeting molecules like Nrf2, NFκβ, E2, E2S, SULT1E1 and MMP expression /activity levels via Oxidative stress. Oxidants/ antioxidants are also regulated via Nrf2 and E2. Based on the current studies, we have made an attempt to sort out or cleave this mechanism in two different pathways which can be utilized for prevention and treatment (Fig-3).

## Data Availability

Yes available

## Acknowledgements

University Grants Commission, New Delhi provided JRF and SRF to AN who is a Ph.D. students working in the Post Graduate Department of Biochemistry, OIST.

## Compliance with Ethical Standards

### Funding

Institutional

### Ethical approval

This article does not contain any studies with human participants or animals performed by any of the authors.

### Informed consent

NA

## References

Abdallah MA, Abdullah HI, Kang S, Taylor DD, Nakajima ST, Gercel-Taylor C. Effects of the components of hormone therapy on matrix metalloproteinases in breast-cancer cells: an in vitro study. FertilSteril. 2007; 87(4):978–981.

Bernstein L, Ross RK. Endogenous hormones and breast cancer risk. Epidemiol Rev. 1993; 15(1): 48–65.

Cai, D., Yin, S., Yang, J., Jiang, Q., and Cao, W. (2015). Histone deacetylase inhibition activates Nrf2 and protects against osteoarthritis. Arthritis Res. Ther.17:269.

Chang J, Zhang Y, Li Y, Lu K, Shen Y, Guo Y, Qi Q, Wang M, Zhang S.NrF2/ARE and NF-κB pathway regulation may be the mechanism for lutein inhibition of human breast cancer cell. Future Oncol. 2018; 14(8):719-726.

Chetrite GS (1), Ebert C, Wright F, Philippe AC, Pasqualini JR. Control of sulfatase and sulfotransferase activities by medrogestone in the Hormone-dependent MCF-7 and T-47D human breast cancer cell lines. J Steroid BiochemMol Biol. 1999; 70(1-3):39–45.

Chia CY, Kumari U and Casey PJ: Breast cancer cell invasion mediated by Gα12 signaling involves expression of interleukins-6 and −8, and matrix metalloproteinase-2. J Mol Signal. 2014; 9: 6.

Duffy MJ, Maguire TM, Hill A, McDermott E, O’Higgins N. Metalloproteinases: role in breast carcinogenesis, invasion and metastasis. Breast Cancer Res. 2000; 2(4):252–257.

Egeblad M, Werb Z. New functions for the matrix metalloproteinases in cancer progression.Nat Rev Cancer. 2002; 2(3):161–174.

Fang J, Shing Y, Wiederschain D, Yan L, Butterfield C, Jackson G, Harper J, Tamvakopoulos G, Moses MA. Matrix metalloproteinase-2 is required for the switch to the angiogenic phenotype in a tumor model. ProcNatlAcadSci U S A. 2000; 97(8):3884–3889.

Gialeli C, Theocharis AD, Karamanos NK. Roles of matrix metalloproteinases in cancer progression and their pharmacological targeting. FEBS J. 2011; 278(1):16–27. [PubMed: 21087457]

Hayes JD, McMahon M. NRF2 and KEAP1 mutations: permanent activation of an adaptive response in cancer. Trends Biochem Sci. 2009; 34(4):176–88

Hudelist G(1), Wülfing P, Kersting C, Burger H, Mattsson B, Czerwenka K, Kubista E, Gschwantler-Kaulich D, Fink-Retter A, Singer CF.Expression of aromatase and estrogen sulfotransferase in preinvasive and invasive breast cancer. J Cancer Res ClinOncol. 2008; 134(1):67-73. 8.

Iochmann S, Bléchet C, Chabot V, Saulnier A, Amini A, Gaud G, Gruel Y and Reverdiau P: Transient RNA silencing of tissue factor pathway inhibitor-2 modulates lung cancer cell invasion. ClinExp Metastasis. 2009; 26: 457–467.

Johanna M. Pellikainen, Kirsi M. Ropponen, Vesa V. Kataja, Jari K. Kellokoski, Matti J. Eskelinen, and Veli-Matti Kosma. Expression of Matrix Metalloproteinase (MMP)-2 and MMP-9 in Breast Cancer with a Special Reference to Activator Protein-2, HER2, and Prognosis.Clinical Cancer Research.2004; 10: 7621–7628.

Karin M, Lin A: NF-kappa B at the crossroads of life and death. Nat Immunol. 2002;3: 221–227.

Katayama H, Paczesny S, Prentice R, Aragaki A, Faca VM, Pitteri SJ, Zhang Q, Wang H, Silva M,Kennedy J, Rossouw J, Jackson R, Hsia J, Chlebowski R, Manson J, Hanash S. Application of serum proteomics to the Women’s Health Initiative conjugated equine estrogens trial reveals a multitude of effects relevant to clinical findings. Genome Med. 2009; 1(4):47.

Katunina AI, Gershtein ES, Ermilova VD, Tereshkina IV, Nazarenko AY, Tyleuova AA, Dvorova EK, Karabekova ZK, Gritskevich MV, Berezov TT. Matrix metalloproteinases 2, 7, and 9 in tumors and sera of patients with breast cancer. Bull ExpBiol Med. 2011;151(3):359–362.

Khan, N. M., and Haqqi, T. M. (2018). Epigenetics in osteoarthritis: potential of HDAC inhibitors as therapeutics. Pharmacol. Res. 128, 73–79.

Klein T, Bischoff R. Physiology and pathophysiology of matrix metalloproteases. Amino Acids.2010.

Larissa Staurengo-Ferrari, Stephanie Badaro-Garcia, Miriam S N Hohmann, Marília F Manchope, Tiago H Zaninelli, Rubia Casagrande, Waldiceu A Verri Jr. Contribution of Nrf2 Modulation to the Mechanism of Action of Analgesic and Anti-inflammatory Drugs in Preclinical and Clinical Stages. Front Pharmacol. 2019; 9: 1536.

Levi E, Fridman R, Miao HQ, Ma YS, Yayon A, Vlodavsky I. Matrix metalloproteinase 2 releases active soluble ectodomain of fibroblast growth factor receptor 1. ProcNatlAcadSci U S A. 1996; 93(14):7069–7074.

Liotta LA, Tryggvason K, Garbisa S, Hart I, Foltz CM, Shafie S. Metastatic potential correlates with enzymatic degradation of basement membrane collagen. Nature. 1980; 284(5751):67–68.

Liu B, Sun L(1), Liu Q(2), Gong C(2), Yao Y(2), Lv X(2), Lin L(3), Yao H(2), Su F(2), Li D(4), Zeng M(5), Song E (6). A cytoplasmic NF-κB interacting long noncoding RNA blocks IκB phosphorylation and suppresses breast cancer metastasis. Cancer Cell. 2015; 27(3):370–81.

Maiti S, Nazmeen A. Impaired redox regulation of estrogen metabolizing proteins is important determinant of human breast cancers. Cancer Cell Int. 2019;19:111. Published 2019 May 15. doi:10.1186/s12935-019-0826-x

Maiti S, Zhang J, Chen G Redox regulation of human estrogen sulfotransferase (hSULT1E1). BiochemPharmacol. 2007 May 1;73(9):1474–81.

Nazmeen A, Chen G, Ghosh TK, Maiti S. Breast cancer pathogenesis is linked to the intratumoral estrogen sulfotransferase (hSULT1E1) expressions regulated by cellular redox dependent Nrf-2/NFκβ interplay. Cancer Cell Int. 2020 Mar 4;20:70.

Nazmeen A, Maiti S. Oxidant stress induction and signalling in xenografted (human breast cancer-tissues) plus estradiol treated or N-ethyl-N-nitrosourea treated female rats via altered estrogen sulfotransferase (rSULT1E1) expressions and SOD1/catalase regulations. Mol Biol Rep. 2018;45(6):2571–2584. doi:10.1007/s11033-018-4425-z

Nazmeen A, Maiti S, Mandal K, et al. Better Predictive Value of Cancer Antigen125 (CA125) as Biomarker in Ovary and Breast Tumors and its Correlation with the Histopathological Type/Grade of the Disease. Med Chem. 2017;13(8):796–804. doi:10.2174/157340641366617042415545

Nilsson UW, Garvin S, Dabrosin C. MMP-2 and MMP-9 activity is regulated by estradiol and tamoxifen in cultured human breast cancer cells. Breast Cancer Res Treat. 2007; 102(3):253–261.

Patel S, Sumitra G, Koner BC, Saxena A. Role of serum matrix metalloproteinase-2 and −9 to predict breast cancer progression. ClinBiochem. 2011;44(10-11):869–872.

Philips N, McFadden K. Inhibition of transforming growth factor-beta and matrix metalloproteinases by estrogen and prolactin in breast cancer cells.Cancer Lett. 2004; 206(1):63–68.

Pitteri SJ, Hanash SM, Aragaki A, Amon LM, Chen L, BusaldBuson T, Paczesny S, Katayama H, Wang H, Johnson MM, Zhang Q, McIntosh M, Wang P, Kooperberg C, Rossouw JE, Jackson RD, Manson JE, Hsia J, Liu S, Martin L, Prentice RL. Postmenopausal estrogen and progestin effects on the serum proteome. Genome Med. 2009; 1(12):121.

Safranek J, Pesta M, Holubec L, Kulda V, Dreslerova J, Vrzalova J, Topolcan O, Pesek M, Finek J and Treska V: Expression of MMP-7, MMP-9, TIMP-1 and TIMP-2 mRNA in lung tissue of patients with non-small cell lung cancer (NSCLC) and benign pulmonary disease. Anticancer Res 2009; 29: 2513–2517.

Sarah J Nyante, Tengteng Wang, Xianming Tan, Emily F Ozdowski, Thomas J Lawton. Quantitative Expression of MMPs 2, 9, 14, and Collagen IV in LCIS and Paired Normal Breast Tissue.Sci Rep. 2019; 9 (1): 13432.

Sheen-Chen SM, Chen HS, Eng HL, Sheen CC, Chen WJ. Serum levels of matrix metalloproteinase 2 in patients with breast cancer. Cancer Lett. 2001;173(1):79–82.

Shostak K (1), Chariot A. NF-κB, stem cells and breast cancer: the links get stronger. Breast Cancer Res. 2011 Jul 26; 13(4):214.

Shun Kobayashi, Noriko Nakajima, Yoko Ito, Mitsuhiko Moriyama. Effects of Lansoprazole on the Expression of VEGF and Cellular Proliferation in a Rat Model of Acetic Acid-Induced Gastric Ulcer.J Gastroenterol. 2010; 45 (8): 846–58.

Smith SM (1), Lyu YL(2), Cai L (3). NF-κB affects proliferation and invasiveness of breast cancer cells by regulating CD44 expression. PLoS One. 2014;9(9):e106966.

Somiari SB, Somiari RI, Heckman CM, Olsen CH, Jordan RM, Russell SJ, Shriver CD. Circulating MMP2 and MMP9 in breast cancer -- potential role in classification of patients into low risk, high risk, benign disease and breast cancer categories. Int J Cancer. 2006;119(6):1403–1411.

Stoeltzing O, Ahmad SA, Liu W, McCarty MF, Wey JS, Parikh AA, Fan F, Reinmuth N, Kawaguchi M, Bucana CD, et al: Angiopoietin-1 inhibits vascular permeability, angiogenesis, and growth of hepatic colon cancer tumors. Cancer Res. 2003; 63: 3370–3377.

Wang W, Nag SA, Zhang R(1). Targeting the NFκB signaling pathways for breast cancer prevention and therapy.Curr Med Chem. 2015;22(2):264–89.

Wu T, Harder BG, Wong PK, Lang JE, Zhang DD.Oxidative stress, mammospheres and Nrf2-new implication for breast cancer therapy? MolCarcinog. 2015;54(11):1494–502.

Zhang W, Wang F, Xu P, Miao C, Zeng X, Cui X, Lu C, Xie H, Yin H, Chen F, et al: Perfluorooctanoic acid stimulates breast cancer cells invasion and up-regulates matrix metalloproteinase-2/-9 expression mediated by activating NF-κB. ToxicolLett.2014; 229: 118–125.

Zubair H Aghai, Sanjay Kumar, Sabeena Farhath, Mary Ann Kumar, Judy Saslow, Tarek Nakhla, Riva Eydelman, Louise Strande, Gary Stahl, Charles Hewitt, Mirjana Nesin, Irfan Rahman. Dexamethasone Suppresses Expression of Nuclear Factor-kappaB in the Cells of Tracheobronchial Lavage Fluid in Premature Neonates With Respiratory Distress.Pediatr Res. 2006; 59 (6): 811–5.

